# Circulating MicroRNAs in Patients with Vulvar Squamous Cell Carcinoma and its Precursors

**DOI:** 10.1101/2024.12.10.24318763

**Authors:** Julia Rymuza, Angelika Długosz, Kamil Zalewski, Artur Kowalik, Mateusz Bujko, Magdalena Kowalewska

**Author notes:** (JR); (KZ); (MB).

## Abstract

**Background/Objectives:** Vulvar squamous cell carcinoma (VSCC) is a rare gynecologic malignancy, with most cases arising from differentiated vulvar intraepithelial neoplasia (dVIN). Approximately one-third of VSCC cases originate from high-grade squamous intraepithelial lesions (HSIL), which are associated with persistent infection by high-risk human papillomavirus (hrHPV) types. This study aimed to quantify circulating microRNAs (miRNAs) in the plasma of patients with premalignant conditions (dVIN and HSIL) and VSCC using TaqMan Low-Density Arrays.

**Methods:** Plasma samples were collected from 40 patients, including those treated for HSIL, dVIN, and VSCC. Quantitative real-time PCR (qRT-PCR) identified circulating miRNAs differentially expressed in the plasma of VSCC patients compared to those with precancerous lesions.

**Results:** A total of 31 differentially expressed miRNAs (DEMs) were found to be significantly upregulated in plasma from VSCC patients compared to precancerous cases. None of the analyzed miRNAs were able to distinguish VSCC cases based on hrHPV tumor status.

**Conclusions:** This study provides strong evidence that a distinct set of miRNAs can differentiate between plasma samples from VSCC patients and those with precancerous lesions. Thus, these DEMs have potential diagnostic and prognostic value. “Predisposing” DEMs could be developed as biomarkers to aid in the assessment of vulvar lesions, helping to exclude or confirm progression toward cancer.

## 1. Introduction

Vulvar carcinoma, a rare malignancy, accounts for 3 to 5% of gynecologic cancers. Vulvar squamous cell carcinoma (VSCC) is the most common type comprising approximately 90% of malignant tumors in this location. Most vulvar carcinomas arise from differentiated vulvar intraepithelial neoplasia (dVIN) and follow an independent of the human papillomavirus (HPV) pathway, with TP53 mutations being a hallmark of this pathway. Approximately one third of vulvar carcinomas arise from high-grade squamous intraepithelial lesions (HSIL), both associated with persistent infection and with high-risk HPV (hrHPV) types, primarily HPV 16 and 18. The rearrest subtype are HPV-negative and wild-type TP53 VSCCs with the suspected precursor termed verruciform acanthotic VIN verruciform acanthotic VIN (vaVIN) or vulvar aberrant maturation (VAM) [1, 2]. Mutations in TP53 result in aberrant protein expression patterns, as assessed by immunohistochemistry (IHC) with the best-known overexpression of TP53. Most studies report that HPV negativity and abnormal expression of TP53 are associated with worse outcomes in patients with VSCC [1]. WHO recommends documentation the type of VSCC (HPV-associated or HPV-independent) on the pathology reports [2], while the International Federation of Gynecology and Obstetrics (FIGO) advocates that all vulvar cancers be tested for HPV and p16 [3]. p16 IHC is a surrogate marker for hrHPV infection, showing a good correlation with HPV testing [1], and the International Collaboration on Cancer Reporting recommends p16 IHC for an accurate classification of VSCC into the two main groups: HPV-associated and HPV-independent VSCC [4]. However, patients with these etiopathogenic VSCC types are currently treated in the same way.

Similarly to other rare cancers, understanding of the biology of VSCC remains limited [5]. In this context, in the past decade research on the role of microRNAs (miRNAs) in the biology of vulvar carcinoma has been undertaken. The initial study by de Melo Maia et al. [6] identified miRNAs with altered expression patterns in VSCC tumors. Some of these alterations were correlated with clinicopathological characteristics such as lymph node metastases (downregulation of miR-223-5p and miR-19-b1-5p), vascular invasion (downregulation of miR-100-3p and miR-19-b1-5p), HPV infection (upregulation of miR-1274b and downregulation of miR-519b), or advanced disease stage (miR-519b and miR-133a overexpression) in patients with VSCC. The same group [7] demonstrated an association of miR-20a and miR-106a upregulation with deeper tumor invasion in VSCC. Up-regulation of another miR, miR-590-5p, was documented to be associated with lymphatic metastases [8]. Recently, a predisposing miRNA profile consisting of 21 miRNAs was established to predict the risk of developing VSCC on a background of vulvar lichen sclerosus, a chronic inflammatory disorder of unknown etiology [8]. These studies confirm that dysregulation of miRNA expression in tumors is implicated in vulvar carcinogenesis and shed light on their potential functions as oncogenes or tumor suppressors in VSCC.

To date, miRNAs hold promise as tumor biomarkers detectable in blood in a non-invasive liquid biopsy approach. To our knowledge, our study on miR-431-5p is the first study to report on miRNA levels in the blood of patients with VSCC. We found that miR-431-5p levels are increased in plasma from patients with VSCC compared to those with vulvar precancers. Low levels of circulating miR-431-5p were found to be indicative of unfavorable survival in patients with VSCC [9]. In this study, we aimed to quantify circulating microRNA profile in plasma samples of VSCC patients using TaqMan Low Density Arrays.

## 2. Results

Based on the Ct results obtained for the three endogenous controls - U6 snRNA, RNU44, RNU48 - included in the TaqMan MicroRNA Arrays, U6 snRNA was selected for qPCR data normalization. RNU44 was found not to be present in all the analyzed samples while the detection of RNU48 was inconsistent across all the cards and samples, and thus these two molecules were excluded from further analysis. The results were normalized with U6 snRNA expression. MiRs that were absent from all the analyzed samples were excluded, leaving 116 miRNAs for further assessment. Samples without U6 snRNA expression were excluded from analysis. Finally, the discrimination abilities of the 116 miRNAs that passed filtering were assessed for their discriminative abilities between ten precancer samples (5 HSIL and 5 dVIN) and 27 VSCC samples (12 hrHPV-negative and 15 hrHPV-positive).

Data comparison between precancer and VSCC groups revealed 31 differentially expressed miRNAs (DEMs) with statistical significance. A volcano graph for these DEMs, showing distribution of significance [-log10(p-value)] vs. fold change [log2(fold change)] demonstrating 31 miRNAs significantly differentially expressed between the precancer group and the VSCC group (p<0.05) is presented in Figure 1a. The heatmap and hierarchical clustering of significant DEMs are shown in Figure 1b. Normalized levels of miRNA were used to hierarchically cluster samples. We observed samples from patients with precancers (dVIN or HSIL) clustered closer together and the VSCC group quite well separated from the precancer group. None of the analyzed miRNAs was found to differentiate samples from patients with VSCC based on tumor hrHPV status (data not shown).

**Figure 1.**
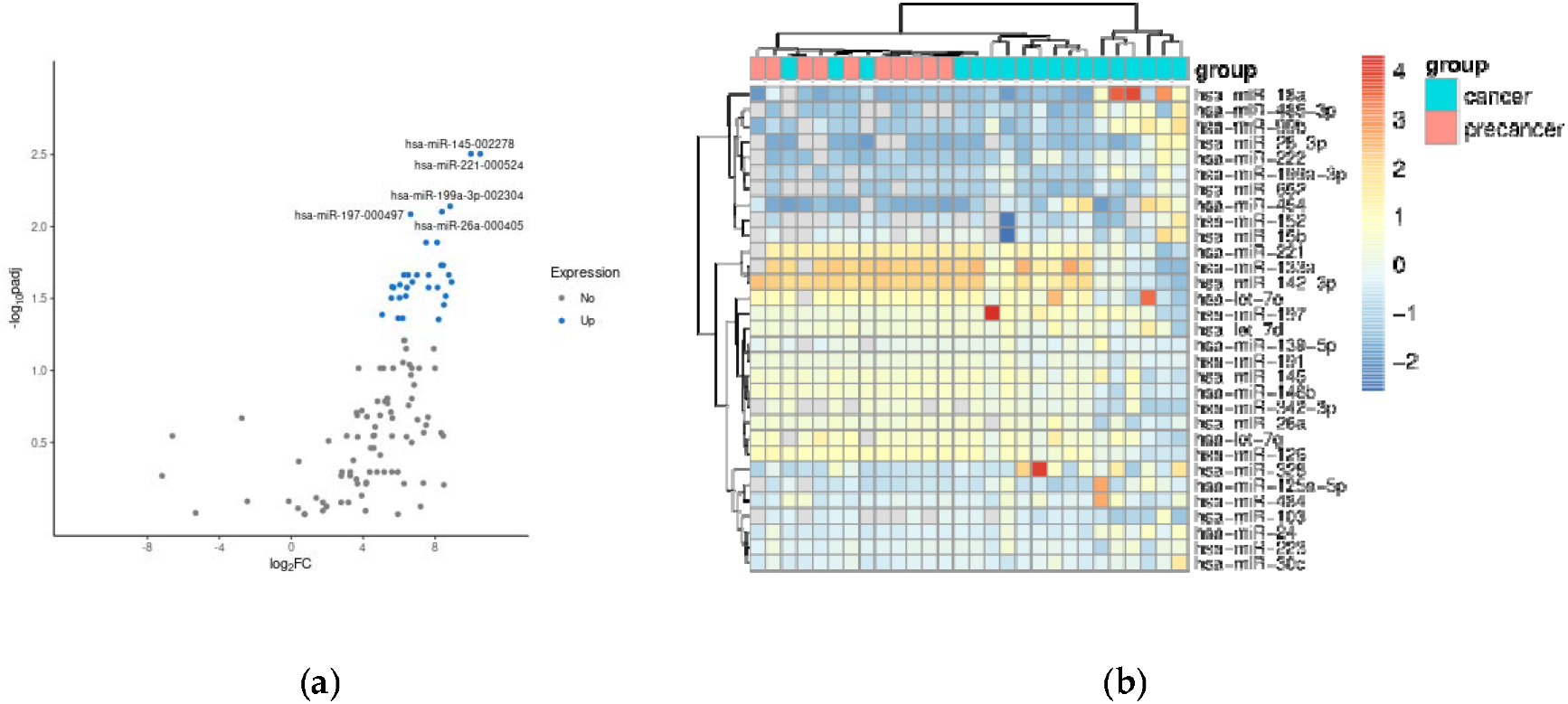
Volcano plot (a) and heatmap (b) for differentially expressed miRNAs between precancer and VSCC groups.

The significant DEMs are highlighted with blue dots, and the top five upregulated are marked with miR symbols. (B) Heat map of significant differentially expressed miRNAs. Heatmap shows the signal of 31 miRNAs (p-value <0.05).

The ROC (Receiver Operating Characteristic) curves for 31 DEMs were generated in order to examine their diagnostic accuracies. The log-rank and ROC AUC tests were performed to assess differences in DEM levels in the two sample groups (plasmas obtained from patients with vulvar precancer vs cancer) and discriminating ability of each miRNA. The ROC curves for DEMs with AUC values higher than 0.9 are shown in Figure 2. The logarithm of fold changes, adjusted p-values for Mann-Whiteny test, AUC values from the ROC tests for DEMs are presented in Table 1.

**Table 1.** Quantitative and statistical measures of 31 high-confidence miRNAs.

| miRNA | log2FC | padj | AUC |
| --- | --- | --- | --- |
| hsa-miR-145-5p | 9.99 | 0.0031 | 0.99 |
| hsa-miR-221-3p | 10.51 | 0.0031 | 0.99 |
| hsa-miR-152 | 8.44 | 0.0186 | 0.98 |
| hsa-miR-199a-3p | 8.83 | 0.0072 | 0.94 |
| hsa-miR-342-3p | 8.33 | 0.0186 | 0.94 |
| hsa-miR-26a | 8.38 | 0.0079 | 0.93 |
| hsa-miR-197 | 6.64 | 0.0082 | 0.92 |
| hsa-miR-485-3p | 7.64 | 0.0217 | 0.91 |
| hsa-miR-28-3p | 8.14 | 0.0265 | 0.91 |
| hsa-miR-15b | 8.92 | 0.0243 | 0.90 |
| hsa-miR-133a | 8.11 | 0.0129 | 0.90 |
| hsa-miR-191 | 7.50 | 0.0129 | 0.89 |
| hsa-miR-652 | 8.59 | 0.0305 | 0.89 |
| hsa-miR-103 | 8.49 | 0.0350 | 0.89 |
| hsa-miR-139-5p | 6.50 | 0.0217 | 0.87 |
| hsa-let-7e | 8.76 | 0.0217 | 0.86 |
| hsa-miR-126 | 6.99 | 0.0217 | 0.86 |
| hsa-miR-30c | 6.26 | 0.0217 | 0.86 |
| hsa-let-7d | 6.73 | 0.0243 | 0.84 |
| hsa-let-7g | 5.60 | 0.0263 | 0.84 |
| hsa-miR-484 | 6.04 | 0.0254 | 0.84 |
| hsa-miR-222 | 6.37 | 0.0305 | 0.83 |
| hsa-miR-99b | 5.57 | 0.0314 | 0.83 |
| hsa-miR-146b | 7.63 | 0.0265 | 0.83 |
| hsa-miR-223 | 6.43 | 0.0265 | 0.83 |
| hsa-miR-328 | 5.69 | 0.0265 | 0.83 |
| hsa-miR-24 | 6.02 | 0.0313 | 0.82 |
| hsa-miR-454 | 5.05 | 0.0410 | 0.81 |
| hsa-miR-125a-5p | 6.19 | 0.0434 | 0.81 |
| hsa-miR-18a | 5.95 | 0.0434 | 0.80 |
| hsa-miR-142-3p | 8.20 | 0.0443 | 0.79 |

Abbreviations: log2 FC: Log2 fold-change; padj: adjusted p-value; AUC: Area Under the ROC curve.

**Figure 2.**
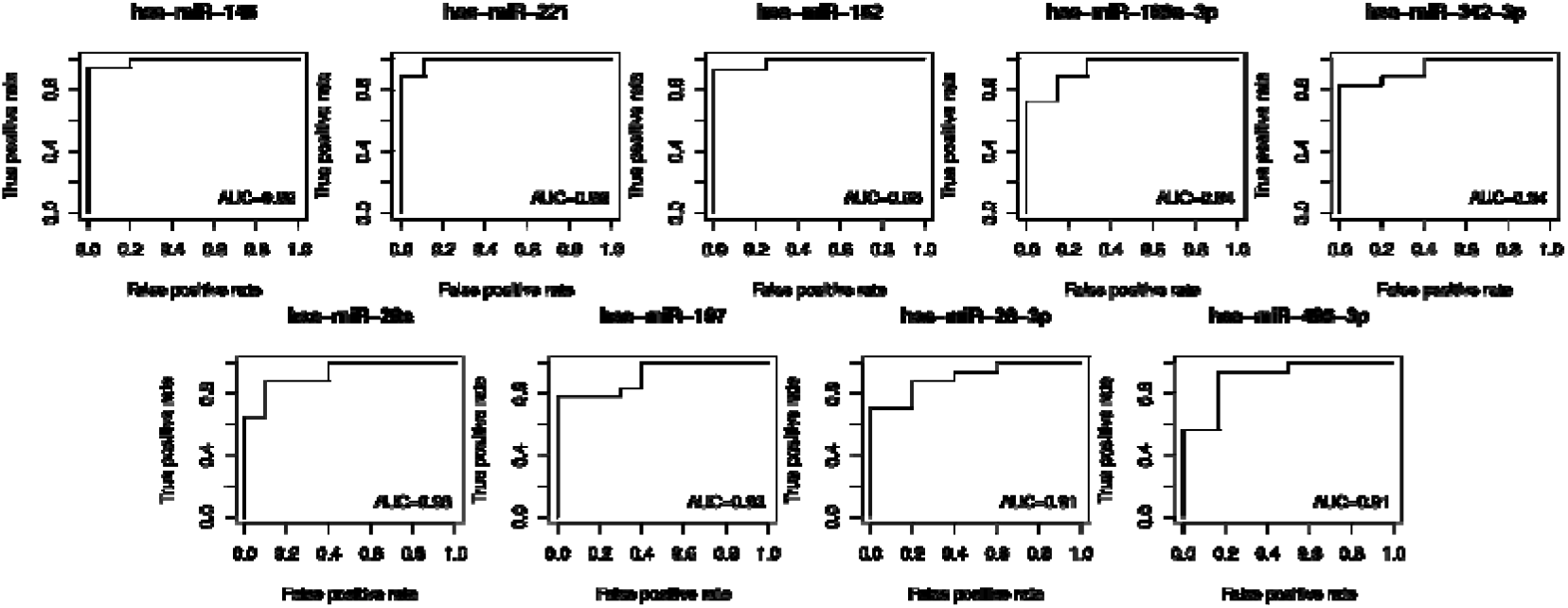
Receiver operating characteristic curves for discrimination of vulvar precancers and cancer based on the plasma levels of the top DEMs (>0.9 AUC).

The significance of differences in levels of DEMs between the three sample groups, i.e. plasma samples from patients with HSIL, dVIN and VSCC was examined with the Kruskal–Wallis test and the post-hoc Dunn’s multiple comparison. The results for seven miRs of the best discriminatory performance for HSIL, dVIN and VSCC samples are depicted in Figure 3. Of note, in this small sample group, miR-222 levels in plasma samples were differentiating between patients with HSIL and dVIN.

**Figure 3.**
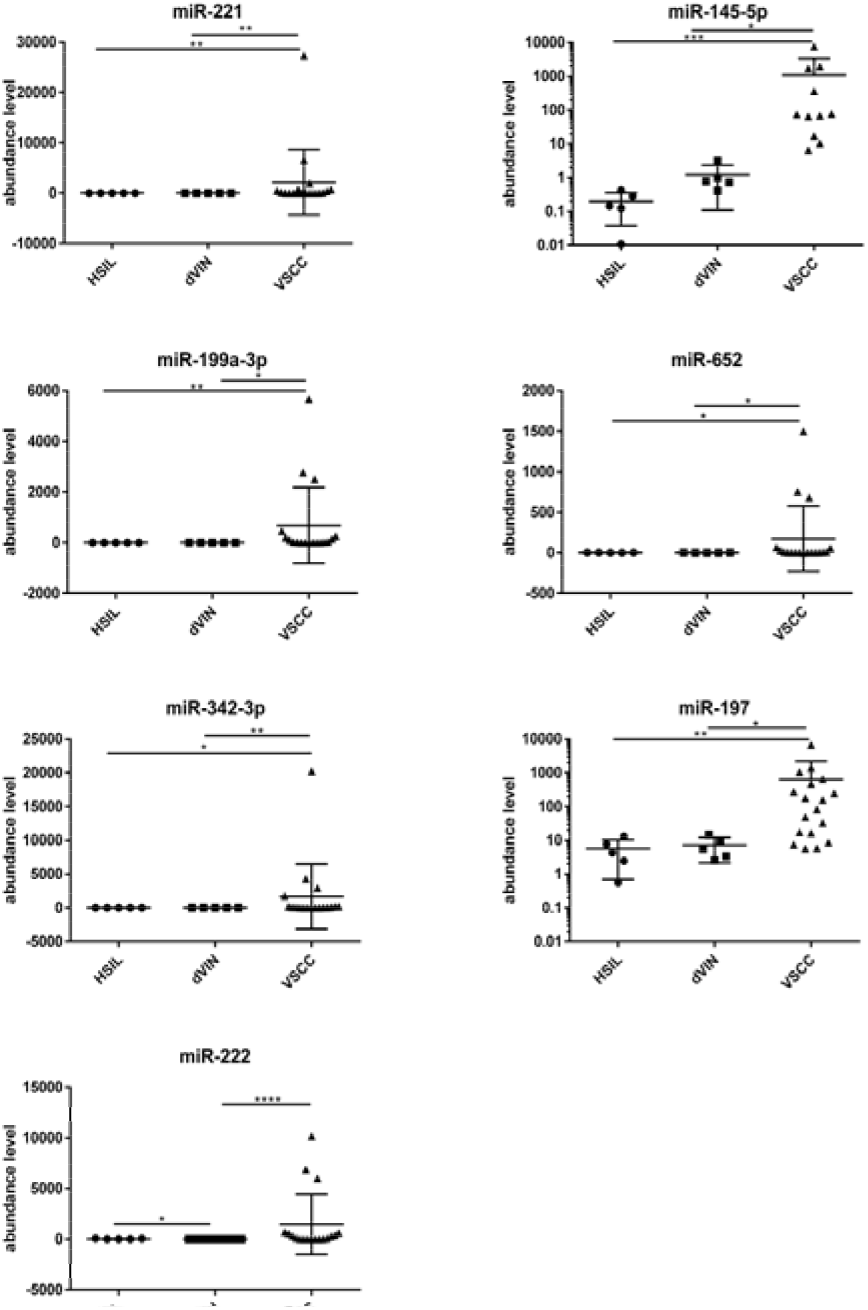
Levels of seven selected miRNAs (mean ± SD) in plasma samples of patients with vulvar precancers and cancers. Significant alternations are indicated by asterisks (*p-value, ≤ 0.05; **, p-value ≤ 0.01; *** p-value ≤ 0.001; ****, p-value ≤ 0.0001).

## 3. Discussion

MiRNAs, small noncoding RNA molecules that influence various cellular processes by modulating the expression of target genes. In cancer, miRNAs can function as oncogenes (oncomiRs) or tumor suppressors, and their aberrant expression profiles have been associated with various types of tumors, including female genital tract tumors [12]. In vulvar carcinoma, specific miRNAs have been associated with tumorigenesis. Some preliminary results of in vitro experiments suggested miRNA-based therapeutic option against VSCC [7]. However, these preliminary data require further verification and more information on the role of specific miRs in vulvar carcinogenesis. For example, downregulation of miR-223-5p was found to be correlated with lymph node metastases in VSCC patients [6] while another study [13] described miR-223-5p as an oncomiR in vulvar carcinoma providing a putative mechanism of its function in tumor invasion.

Currently, information regarding circulating miRNAs in VSCC is limited. Yet, these molecules hold potential as diagnostic biomarkers, particularly in the context of liquid biopsy, where their relative stability in biological fluids allows for non-invasive detection and monitoring of tumor-specific molecular changes. Previously, our group demonstrated that lowered levels of circulating miR-431-5p are associated with poor prognosis in patients with VSCC, supporting the potential utility examination of miRs in the blood to identify patients at high risk of progression [9]. By analyzing miRNAs abundance patterns in blood of patients treated for HSIL, dVIN and VSCC, this study aimed to identify circulating miRNAs with qRT-PCR that may aid in distinguishing between malignant and premalignant conditions by plasma analysis. Our analysis demonstrated differential levels of circulating miRNAs between patients with vulvar premalignant lesions and carcinoma. Our study revealed that among the analyzed miR, hsa-miR-221-3p and hsa-miR-145-5p were highly upregulated in plasmas of the VSCC group compared to the premalignant group.

The top among up-regulated DEMs in a set of genital tumors, miR-221 is an extensively investigated oncomiR which plays an important role in the progression of many malignancies [10]. Of interest, apart from targeting mRNAs involved in main survival pathways, via silencing inflammatory genes it can induce macrophage tolerization [11]. miR-221 was shown to target TP53BP2, a positive regulator of the TP53 pathway [12]. miR-221 has a potential as a diagnostic and prognostic biomarker, with its detection in body fluids offering a promising liquid biopsy tool. It is also a therapeutic target of various reported inhibition strategies, including the locked nucleic acid (LNA) inhibitor examined in an in-human study [13]. miR-221 is encoded by one miR-221/222 gene cluster with miR-222, another DEMs identified in our study. Interestingly, miR-222 levels in plasma samples were revealed to differentiate patients with HSIL and dVIN. This finding promising in the context of diagnostic value of miR-222, although statistically significant, should be interpreted with caution due to small sample numbers examined.

On the contrary, the second of the top up-regulated DEMs in the blood of VSCC patients, miR-145, is well-established tumor suppressor [14]. Wide range of cancers are characterized by reduced levels of this microRNA. It was demonstrated that miR-145-5p can regulate cell proliferation and migration by modulating various signaling pathways, such as MAPK and PI3K/AKT [15]. Due to the anti-oncogenic effects of miR-145, the therapeutic approaches proposed in oncology involve restoring its expression or using its mimics. Given its identified role of miR-145 as a tumor suppressor, our finding of its upregulations seems unexpected. Furthermore, miR-145 was shown to suppress ovarian cancer cell invasion and migration through targeting i.a. HMGA2 (High Mobility Group AT-Hook 2) gene [16]. Our group has previously documented increase HMGA2 protein abundances in VSCC tumors, as compared to vulvar premalignant lesions [17]. However, in the blood of cancer patients miR-145 levels can be significantly reduced, as recently reported for head and neck squamous cell carcinoma [18]. The mechanistic biological explanation for the high increase of miR-145 in blood of VSCC patients remains to be solved. The same notion is true for all identified DEMs that can be used to discriminate patients with VSCC from patients with precursor lesions. Our data provide preliminary information on the detection of miRNAs that are potentially involved in early vulvar carcinogenesis processes.

## 4. Materials and Methods

### Patients

Serum samples were obtained from 40 patients treated for premalignant vulvar lesions (HSIL, n = 7 and dVIN, n = 6) and VSCC (n = 27) at the Maria Sklodowska-Curie National Research Institute of Oncology Warsaw, Poland and at the Holycross Cancer Center in Kielce, Poland, between December 2004 and March 2017. The cancer patients enrolled in the study had microscopically confirmed primary VSCC in the early stages (FIGO stage I, n=18) and advanced stages (FIGO stage III, n=9), aged 46 to 83 (median age – 68).

### HPV Genotyping

The HPV status of precancer lesions and vulvar malignant tumors was determined as described previously [19] using the AmpliSens HPV HCR-genotype-titre-FRT test (InterLabService Ltd.).

### RNA Isolation from plasma and qPCR

Plasma samples obtained from patients and healthy donors were banked at −70 ° C before their analysis. Total RNA was isolated from 200 µL of plasma using the miRNeasy Serum/Plasma Kit (Qiagen). 100 ng of total RNA were used for miRNA reverse transcription using Pool A of Megaplex™ RT Human Primers (Applied Biosystems) in the Veriti™ 96 Well Thermal Cycler (Applied Biosystems). The miRNA cDNA was then preamplified using Megaplex™ PreAmp Primers, Pool A, in the Gene Amp® PCR System 9700 (Applied Biosystems). Following dilution, preamplified microRNA cDNA was loaded onto TaqMan® MicroRNA Arrays A v2.0 (Applied Biosystems) containing 384 TaqMan® MicroRNA Assays per card. The qRT-PCR amplification and data acquisition were performed on the ViiA™ 7 Real-Time PCR System (Thermo Fisher Scientific|Applied Biosystems). These microfluidic cards allow the quantification of 377 human microRNAs. The reverse transcription, pre-amplification, dilution, and qPCR steps were performed according to the manufacturer’s protocols (Applied Biosystems).

### Data analysis

The collected data were analyzed using threshold-cycle (Ct) values for the miRNAs with the Relative Quantification (RQ) Application Module on the Thermo Fisher Cloud using automated baseline and manually set threshold values. The results were normalized with endogenous control included in the TaqMan MicroRNA Arrays. Ct values were exported and the relative amounts of each miRNA in the plasma of patients were calculated using the 2−ΔCtT method.

### Statistical analyses

The Mann-Whitney test was applied to find differentially expressed miRNAs between two sample groups. miRNA performance as a marker was assessed using the area under the curve of receiver operating characteristic curve (ROC AUC). Pearson correlation was calculated to check association between miRNA expression and HPV status. These analyses were performed using python library scipy [20].

The nonparametric Kruskal–Wallis test with the post-hoc Dunn’s multiple comparison test was used to assess the significance of differences in levels of selected miRs between the three sample groups. This statistical analysis was performed and the results were visualized using GraphPad Prism 6.07 (GraphPad Software, USA). Differences at p < 0.05 were considered significant.

## 5. Conclusions

In summary, despite a small sample size, our study provides a strong indication that there is a set of miRNAs differentiating between plasma from patients with vulvar carcinoma and its precancer lesions. Distinguishing between VSCC and its precancers is critically important for patient management and outcomes. Early and accurate differentiation can prevent overtreatment of benign or precancerous conditions and ensure timely treatment of malignancies. In our study, several circulating miRNAs, particularly hsa-miR-221-3p and hsa-miR-145-5p, showed significantly increased levels in plasma of VSCC patients as compared to plasma samples of women with vulvar premalignant lesions. This finding suggesting potential diagnostic value of these molecules. Furthermore, understanding the molecular differences between these conditions, such as those reflected in circulating biomarkers, such as microRNAs, may improve diagnostic precision and guide personalized therapeutic strategies. It should be further investigated whether the profile of circulating miRs or specific miRs could be useful in predicting the progression of vulvar lesions to VSCC.

## Author Contributions

Conceptualization, M.K.; methodology, J.R., M.B. and M.K.; software, J.R., A.D. and M.K.; investigation, A.D. and M.K.; resources, K.Z. and A.K.; writing—original draft preparation, M.K.; writing— review and editing, all the authors, visualization, J.R. and M.K.; supervision, M.B. and M.K.; project administration, M.K.; funding acquisition, M.K. and K.Z. All authors have read and agreed to the published version of the manuscript.

## Funding

This research was funded by the Polish National Science Centre, grant number 2013/23/N/NZ5/03284.

## Institutional Review Board Statement

The study was conducted according to the guidelines of the Declaration of Helsinki, and approved by the Institutional Review Ethics Committees of the Maria Sklodowska-Curie National Research Institute of Oncology Warsaw, Poland (No. 44/2002, 16/2015) and of the Holycross Cancer Center in Kielce, Poland (No. 15/2014).

## Informed Consent Statement

Informed consent was obtained from all subjects involved in the study.

## Data Availability Statement

Data are available upon reasonable request from the corresponding author.

## Conflicts of Interest

The authors declare no conflict of interest. The funders had no role in the design of the study; in the collection, analyses, or interpretation of data; in the writing of the manuscript, or in the decision to publish the results.

## Notes

### Competing Interest Statement

The authors have declared no competing interest.

